# Chagas disease and SARS-CoV-2 coinfection does not lead to worse in-hospital outcomes: results from the Brazilian COVID-19 Registry

**DOI:** 10.1101/2021.03.22.21254078

**Authors:** Israel Molina, Milena S. Marcolino, Magda C. Pires, Lucas Emanuel F. Ramos, Rafael T. Silva, Milton H. Guimarães, Isaias José R. de Oliveira, Rafael L.R. Carvalho, Aline Gabrielle S. Nunes, Ana Lara R. M. de Barros, Ana Luiza B. A. Scotton, Angélica Aparecida C. Madureira, Bárbara L. Farace, Cíntia A. de Carvalho, Fernanda D. A Rodrigues, Fernando Anschau, Fernando A. Botoni, Guilherme F. Nascimento, Helena Duani, Henrique C. Guimarães, Joice C. de Alvarenga, Leila B. Moreira, Liege B. Zandoná, Luana F. de Almeida, Luana M. Oliveira, Luciane Kopittke, Luís C. Castro, Luisa Elem A. Santos, Máderson A. S. Cabral, Maria Angélica P. Ferreira, Natália C. S. Sampaio, Neimy R. de Oliveira, Saionara C. Francisco, Sofia J. T. S. Lopes, Tatiani O. Fereguetti, Veridiana B. dos Santos, Victor Eliel B. de Carvalho, Yuri C. Ramires, Antonio Luiz P. Ribeiro, Freddy Antonio B. Moscoso, Rogério Moura, Carisi A. Polanczyk, Maria do Carmo P. Nunes

## Abstract

**Objective:** Chagas disease (CD) continues to be a major public health burden in Latina America, where co-infection with SARS-CoV-2 can occur. However, information on the interplay between COVID-19 and Chagas disease is lacking. Our aim was to assess clinical characteristics and in-hospital outcomes of patients with CD and COVID-19, and to compare it to non-CD patients.

**Methods:** Patients with COVID-19 diagnosis were selected from the Brazilian COVID-19 Registry, a prospective multicenter cohort, from March to September, 2020. CD diagnosis was based on hospital record at the time of admission. Study data were collected by trained hospital staff using Research Electronic Data Capture (REDCap) tools. Genetic matching for sex, age, hypertension, DM and hospital was performed in a 4:1 ratio.

**Results:** Of the 7,018 patients who had confirmed infection with SARS-CoV-2 in the registry, 31 patients with CD and 124 matched controls were included. Overall, the median age was 72 (64.-80) years-old and 44.5% were male. At baseline, heart failure (25.8% vs. 9.7%) and atrial fibrillation (29.0% vs. 5.6%) were more frequent in CD patients than in the controls (p<0.05 for both). C-reactive protein levels were lower in CD patients compared with the controls (55.5 [35.7, 85.0] vs. 94.3 [50.7, 167.5] mg/dL). Seventy-two (46.5%) patients required admission to the intensive care unit. In-hospital management, outcomes and complications were similar between the groups.

**Conclusions:** In this large Brazilian COVID-19 Registry, CD patients had a higher prevalence of atrial fibrillation and chronic heart failure compared with non-CD controls, with no differences in-hospital outcomes. The lower C-reactive protein levels in CD patients require further investigation.

**Key messages:** *What is already known about this subject?:*

- Preexisting cardiovascular disease enhances vulnerability to COVID-19.
- Co-infection with SARS-CoV-2 and *T*.*cruzi* can occur in patients living in areas in which both infections are epidemic.

*What does this study add?:*

- Despite a higher frequency of chronic heart failure and atrial fibrillation, our findings do not suggest that co-infection with *T*.*cruzi* and SARS-CoV-2 worsens in-hospital outcomes.
- Chagas disease patients were observed to have lower C-reactive protein (CRP) levels.

*How might this impact on clinical practice?:*

- Given the current circulation of SARS-CoV-2 at high levels and millions of *T* c*ruzi*-infected individuals living in Brazil, the risk for co-infections substantially increases.
- Further studies are needed to investigate why CRP levels were lower in CD patients. We hypothesized that CD patients might have a lower risk of unregulated inflammatory response to COVID-19, as they already have an active chronic inflammatory and immune response response triggered by *T*.*cruzi* infection.

## Introduction

Since the first case of coronavirus disease 19 (COVID-19) described in Brazil on February 26^th^, 2020, SARS-CoV 2 infection has evolved as a global pandemic. The disease has a wide spectrum of clinical manifestations, ranging from asymptomatic cases to severe pneumonia and acute respiratory distress syndrome. [1,2]

Although the great majority of symptoms are unspecified, mild, flu-like or belonging to respiratory sphere, other organs could be affected, as the cardiovascular system. COVID-19 has been associated with multiple cardiac manifestations, including cardiac arrhythmias, myocardial infarction, acute heart failure and acute fulminant myocarditis. Cardiovascular involvement has shown to be associated with increased mortality. [3,4]

Underlying comorbidities have been widely associated with a worse prognosis for COVID-19 patients, since viral infections could act as triggers for worsening of chronic diseases.[5–7] Chagas disease (CD) is a multisystemic disorder, potentially affecting, cardiovascular, digestive, and neurological systems. It is the most common cause of infectious cardiomyopathy worldwide, and it may play a role in the clinical prognosis of COVID-19 patients. [8,9] Although CD is endemic in Latin America, it has been recognized that the disease is now a worldwide concern, as the disease spread with population movements from endemic to non-endemic countries.[10] In Brazil, CD still remains a public health challenge, being one the countries with more absolute number of patients and an annual incidence rate of approximately 0.16 per 100,000 inhabitants/year.[11]

Potential interactions between COVID-19 and Chagas cardiomyopathy could be probable, because both conditions share the same immunological pathway. SARS-CoV-2 spike proteins bind to angiotensin-converting enzyme-2 (ACE-2), which is needed to invade the host cell. On the other hand, ACE2 is involved in heart function and the development of hypertension and diabetes mellitus (DM), risk factors frequently observed in patients with chronic Chagas cardiomyopathy. [12,13] Those patients could have increased levels of ACE2 because of the chronic use of ACE inhibitors and/or angiotensin receptor blockers (ARBs).

Limited information is available regarding the characteristics and outcomes of patients with CD and COVID-19. Therefore, we aim to describe the characteristics, laboratory, and imaging findings, as well as in-hospital outcomes of CD and COVID-19 coinfected patients included in the Brazilian COVID-19 Registry.

## Methods

This manuscript adheres to the Strengthening the Reporting of Observational Studies in Epidemiology (STROBE) guideline. [14]

### Study design and subjects

Patients were selected from the Brazilian COVID-19 Registry, a prospective multicenter cohort project with 37 participant hospitals in 17 cities from three Brazilian states (Minas Gerais, Pernambuco, Rio Grande do Sul, Santa Catarina, São Paulo). Details of the cohort were published elsewhere. [5]

COVID-19 diagnosis was confirmed through real time polymerase-chain reaction (RT-PCR) nasopharyngeal and oropharyngeal swab testing or anti-SARS-CoV-2 IgM detected in serological assay in serum or plasma sample, according to World Health Organization guidance. [15]

For the present study, patients with previous history of CD recorded in the database were selected. CD diagnosis were retrieved by their own hospital record or self-referred by the patient. Patients were admitted from March 1 to September 30, 2020. At the moment of the analysis 7,018 patients were introduced in the registry, 31 of those were classified as suffering from CD.

### Data collection

Study data were collected by trained hospital staff or interns using Research Electronic Data Capture (REDCap) tools. [16] Medical records were reviewed to collect data on patients’ demographic and clinical characteristics, including age, sex, pre-existing medical conditions and home medications; COVID-19 symptoms at hospital presentation; clinical assessment upon hospital admission, third and fifth admission days; laboratory, imaging, electrocardiographic data; inpatient medications, treatment and outcomes. Definitions were published elsewhere. [5]

### Statistical analysis

Genetic matching for sex, age, hypertension, DM and hospital was performed in a 4:1 ratio (MatchIt package in R). Sample size of 132 controls was calculated considering and expected risk ratio for mortality 2.5 in CD-group, power of 80%, alfa-error probability of 5% for a 4:1 CD/control.

Categorical data were presented as absolute numbers and proportions, and continuous variables were expressed as medians and interquartile ranges. The χ2 and Fisher Exact test were used to compare the distribution of categorical variables, and the Wilcoxon-Mann–Whitney test for continuous variables. Results were considered statistically significant if the two-tailed P-value was < 0.05. All statistical analysis was performed with R software (version 4.0.2).

### Ethics

The study was approved by the National Commission for Research Ethics (CAAE 30350820.5.1001.0008). Individual informed consent was waived owing to the pandemic situation and the use of deidentified data, based on medical chart review only.

## Results

### Patient characteristics at hospital admission

From the 155 patients included in the study (Figure 1), 31 were reported as having Chagas disease, and 124 were matched controls. The median age was 72.0 (64.0-79.5) years-old and 44.5% were male. Hypertension (65.8%), DM (32.3%), chronic obstructive pulmonary disease (COPD) in (16.7%), chronic heart failure (12.9%) and atrial fibrillation (10.3%) were the most frequent comorbidites.

**Figure 1.**
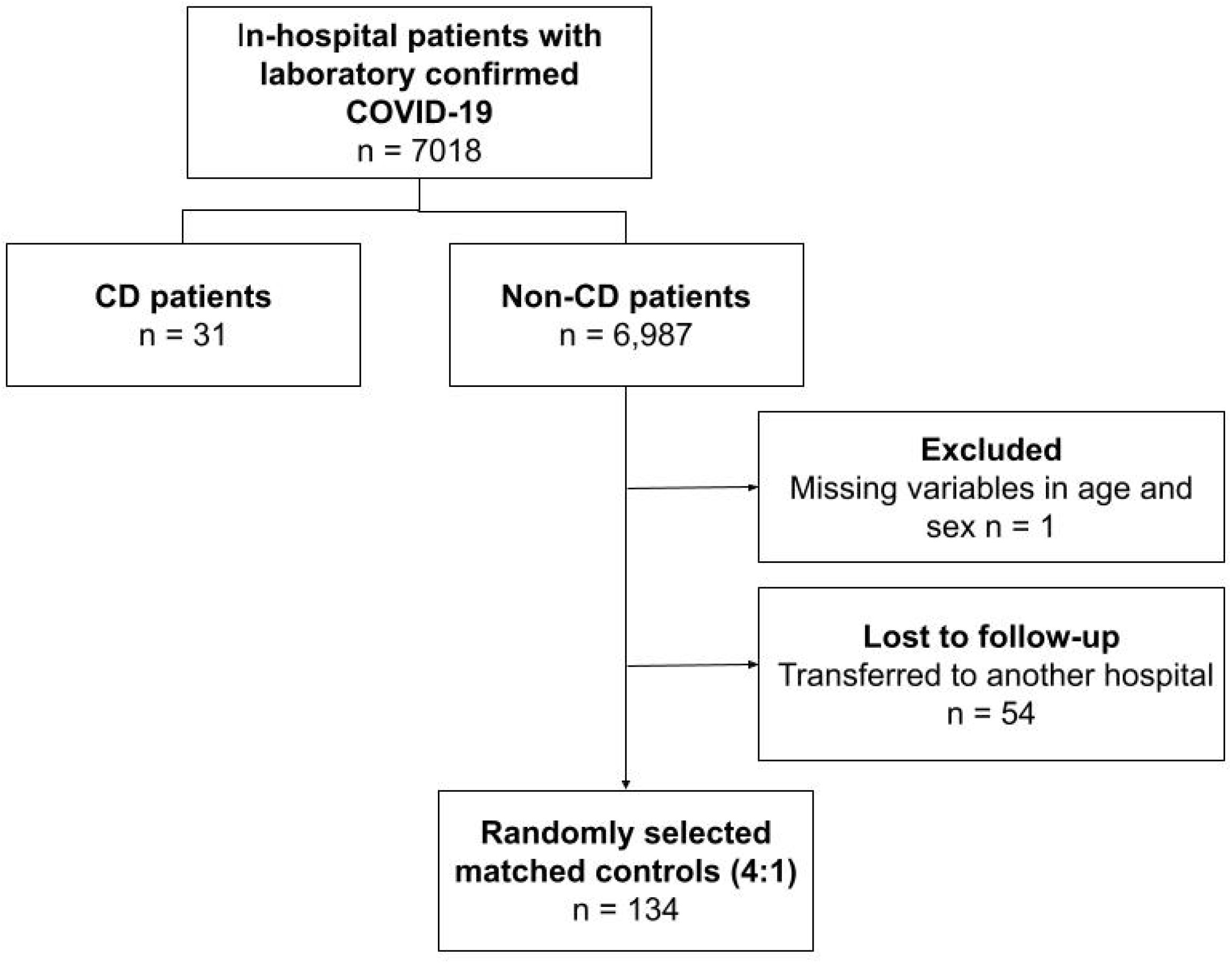
Flowchart of COVID-19 patients included in the study.

Patients were from 11 hospitals, with average 382 beds (ranging from 60 to 936 beds). Nine of them (81.8%) were public, 7 (63.6%) were teaching hospitals and 8 (72.7%) were reference centers for COVID-19 treatment.

When comparing CD patients with controls (Table 1), there were no significant differences in demographic and medical characteristics, except for the prevalence of chronic heart failure (8 [25.8%] vs 12 [9.7%]; p=0.031) and atrial fibrillation (9 [29.0%] vs 7 [5.6%]; p < 0.001), which were more prevalent in CD patients. Although the median number of comorbidities was higher in CD patients (3.0 [2.0, 4.0] vs. 2.0 [1.0, 3.0]), this difference did not reach statistical significance (p=0.119).

**Table 1.** Demographic characteristics and medical history data of the study population at baseline

|  | **CD patients**<br>**(n=31)** | **Controls**<br>**(n=124)** | **p-value** |
| --- | --- | --- | --- |
| **Age\*** (years) | 74.0 (64.5, 79.0) | 72.0 (64.0, 80.0) | 0.856 |
| **Male sex\*** | 14 (45.2%) | 55 (44.4%) | >0.999 |
| **Comorbidities\*\*** |  |  |  |
| Total number |  |  | 0.461 |
| 0 | 3 (9.7%) | 11 (8.9%) |  |
| 1 | 3 (9.7%) | 27 (21.8%) |  |
| 2 | 9 (29.0%) | 39 (31.5%) |  |
| 3 | 7 (22.6%) | 26 (21.0%) |  |
| 4 | 6 (19.4%) | 16 (12.9%) |  |
| ≥ 5 | 3 (9.7%) | 5 (4.0%) |  |
| *Cardiovascular diseases* |  |  |  |
| Hypertension* | 20 (64.5%) | 82 (66.1%) | >0.999 |
| Ischemic cardiopathy | 1 (3.2%) | 6 (4.8%) | >0.999 |
| Chronic heart failure | 8 (25.8%) | 12 (9.7%) | **0.031** |
| Atrial fibrillation/flutter | 9 (29.0%) | 7 (5.6%) | **<0.001** |
| Stroke | 2 (6.5%) | 8 (6.5%) | >0.999 |
| Pacemaker | 1 (3.2%) | 0 (0.0%) | 0.200 |
| *Respiratory diseases* |  |  |  |
| Asthma | 1 (3.2%) | 9 (7.3%) | 0.688 |
| COPD | 8 (25.8%) | 18 (14.5%) | 0.216 |
| *Metabolic diseases* |  |  |  |
| Diabetes mellitus* | 10 (32.3%) | 40 (32.3%) | >0.999 |
| Obesity (BMI>30) | 1 (3.2%) | 10 (8.1%) | 0.695 |
| *Other conditions* |  |  |  |
| Cirrhosis | 0 (0.0%) | 2 (1.6%) | >0.999 |
| Psychiatric condition | 1 (3.2%) | 9 (7.3%) | 0.688 |
| Chronic renal disease | 0 (0.0%) | 3 (2.4%) | >0.999 |
| Dyslipidemia | 0 (0.0%) | 1 (0.8%) | >0.999 |
| HIV | 0 (0.0%) | 2 (1.6%) | >0.999 |
| Neoplasia | 3 (9.7%) | 8 (6.5%) | 0.461 |
| Transplantation | 1 (3.2%) | 3 (2.4%) | >0.999 |
| Dementia | 0 (0.0%) | 1 (0.8%) | >0.999 |
| Epilepsy | 0 (0.0%) | 0 (0.0%) | - |
| **Toxic habits** |  |  |  |
| Alcohol | 1 (3.2%) | 6 (4.8%) | >0.999 |
| Tobacco (active or former) | 7 (22.6%) | 35 (28.2%) | 0.684 |

Numbers are presented as medians (P25-P75) or counts (percentages).
BMI: body mass index; CD: Chagas disease; COPD: chronic obstructive pulmonary disease.
\* Controls were paired for age, sex, hospital, hypertension and diabetes.
\*\* This variable does not include Chagas disease.

The median time since from symptom onset to hospital admission was 6 (8-4) days. Dyspnea and cough (dry or productive) were present in more than one half of patients. There were no differences in the clinical presentation between both groups (Table 2).

**Table 2.** Clinical characteristics of the study population at baseline

|  | CD patients (n=31) |  | Controls (n=124) |  |  |
| --- | --- | --- | --- | --- | --- |
|  | Frequency (%)<br>or median (IQR) | Valid<br>cases | Frequency (%)<br>or median (IQR) | Valid<br>cases | p-value |
| Symptoms |  |  |  |  |  |
| Time from symptom<br>onset | 5.0 (3.0, 7.8) | 30 | 6.0 (3.8, 9.2) | 124 | 0.392 |
| Adynamic | 10 (32.3%) | 31 | 37 (29.8%) | 124 | 0.965 |
| Ageusia | 4 (12.9%) | 31 | 7 (5.6%) | 124 | 0.232 |
| Anosmia | 5 (16.1%) | 31 | 10 (8.1%) | 124 | 0.183 |
| Headache | 7 (22.6%) | 31 | 22 (17.7%) | 124 | 0.719 |
| Rhinorrhea | 4 (12.9%) | 31 | 20 (16.1%) | 124 | 0.786 |
| Diarrhea | 3 (9.7%) | 31 | 18 (14.5%) | 124 | 0.573 |
| Dyspnea | 19 (61.3%) | 31 | 73 (58.9%) | 124 | 0.967 |
| Odynophagia | 14 (45.2%) | 31 | 64 (51.6%) | 124 | 0.659 |
| Fever | 4 (12.9%) | 31 | 17 (13.7%) | 124 | >0.999 |
| Hyporexia | 1 (3.2%) | 31 | 5 (4.0%) | 124 | >0.999 |
| Neurological<br>manifestations | 6 (19.4%) | 31 | 34 (27.4%) | 124 | 0.491 |
| Myalgia | 2 (6.5%) | 31 | 19 (15.3%) | 124 | 0.252 |
| Nausea/vomiting | 7 (22.6%) | 31 | 21 (16.9%) | 124 | 0.639 |
| Productive cough | 18 (58.1%) | 31 | 65 (52.4%) | 124 | 0.717 |
| Dry cough | 1 (3.2%) | 31 | 1 (0.8%) | 124 | 0.361 |
| Clinical assessment |  |  |  |  |  |
| Glasgow <15 | 6 (19.4%) | 31 | 24 (19.4%) | 124 | >0.999 |
| HR | 80.0 (72.0, 86.8) | 30 | 84.0 (77.0, 96.0) | 121 | 0.060 |
| HR ≥ 100 bpm | 4 (12.9%) | 31 | 28 (22.6%) | 124 | 0.346 |
| RR | 22.0 (18.5, 26.0) | 27 | 22.0 (18.0, 25.0) | 115 | 0.748 |
| RR ≥ 24 irpm | 16 (51.6%) | 31 | 56 (45.2%) | 124 | 0.658 |
| Sat O2 | 94.0 (91.0, 96.0) | 29 | 94.0 (90.0, 96.0) | 123 | 0.712 |
| Sat O2 < 90% | 7 (22.6%) | 31 | 28 (22.6%) | 124 | >0.999 |
| SF ratio | 402.4 (300.0, 440.5) | 28 | 395.8 (240.0, 438.1) | 123 | 0.316 |
| Invasive ventilation | 3 (9.7%) | 31 | 13 (10.5%) | 124 | >0.999 |
| SBP ≤ 100 mmHg | 1 (3.2%) | 31 | 11 (8.9%) | 124 | 0.462 |
| Inotropic drugs | 12 (38.7%) | 31 | 45 (36.3%) | 124 | 0.967 |

CD: Chagas disease; HR: hear rate; IQR: interquartile range; RR: respiratory rate; SF ratio: Sat O<sub>2</sub>/FiO<sub>2</sub>; valid cases: non missing cases.

Laboratory and imaging findings are presented in Supplementary Table 1 and 2. Median C-reactive protein was lower in CD patients than the controls (55.5 [35.7, 85.0] vs. 94.3 [50.7,167.5] mg/dL). There was no other clinically relevant difference in laboratory exams between groups.

At admission, diffuse interstitial infiltrate pattern and ground glass opacities were the most prevalent findings in the chest X-ray and chest computer tomography (CT), respectively. No significant differences were found in the frequency of abnormalities and radiological progression in both groups, expect for the frequency of pleural effusion in the follow-up CT, more frequent in CD patients.

Among CD, patients 10 had an EKG performed. Of those, 4 patients had atrial fibrillation and 2 had a pacemaker rhythm, so the proportion of patients with sinus rhythm in controls were significantly higher than in CD patients (68.8% vs 40.0%, p = 0.142) (Table 3).

**Table 3.** Electrocardiographic characteristics of the study population at baseline and new abnormalities at follow-up.

|  | **CD patients**<br>**(n=31)** | **Control patients**<br>**(n=124)** | **p-value** |
| --- | --- | --- | --- |
| **ECG at admission** | 10 (32.3%) | 32 (26.0%) | 0.637 |
| Sinus rhythm | 4 (40.0%) | 22 (68.8%) | 0.142 |
| Atrial fibrillation or flutter | 4 (40.0%) | 7 (21.9%) | 0.410 |
| Pacemaker | 2 (20.0%) | 1 (3.1%) | 0.136 |
| Right bundle branch block | 1 (10.0%) | 4 (12.5%) | >0.999 |
| Left bundle branch block | 2 (20.0%) | 1 (3.1%) | 0.136 |
| Left ventricular hemiblock | 0 (0.0%) | 0 (0.0%) |  |
| **New electrocardiographic abnormalities\*** | **N\*=4 (12.9%)** | **N\*=15 (12.3%)** | **>0.999** |
| Rhythm |  |  |  |
| Atrial fibrillation or flutter | 4 (100.0%) | 6 (40.0%) | 0.087 |
| Pacemaker | 1 (25.0%) | 0 (0.0%) | 0.211 |
| Multifocal atrial rhythm | 0 (0.0%) | 1 (6.7%) | >0.999 |
| Supraventricular tachycardia | 0 (0.0%) | 1 (6.7%) | >0.999 |
| Monomorphic ventricular tachycardia | 0 (0.0%) | 3 (20.0%) | >0.999 |
| Polymorphic ventricular tachycardia | 0 (0.0%) | 1 (6.7%) | >0.999 |
| No new rhythm abnormalities | 0 (0.0%) | 4 (26.7%) | 0.530 |
| New long QTc interval | 1 (25.0%) | 2 (13.3%) | 0.530 |
| None | 2 (50.0%) | 4 (26.7%) | 0.557 |

\* New electrocardiographic abnormalities through in-hospital follow-up, and number of patients in which this outcome was assessed.

### Treatment and clinical outcomes

There were no differences regarding the therapeutic strategy among both groups (Table 4), except for a trend of higher frequency of therapeutic anticoagulation in CD patients (19.3% vs. 10.5%, p=0.206). Twenty-four CD patients (77.4%) and 103 controls (83.0%) received corticosteroids (p=0.448). Dexamethasone was used by 64.5% CD patients and 66.1% controls (p>0.999). Macrolides were prescribed for 77.4% in CD patients and 87.1% controls (p=0.255); chloroquine or hydroxychloroquine in 3.2% and 4.8% (p>0.999). Only one patient received remdesivir.

**Table 4.** Medications.

|  | **CD patients**<br>**(n=31)** | **Controls**<br>**(n=124)** | **p-value** |
| --- | --- | --- | --- |
| Azithromycin | 23 (74.2%) | 91 (73.4%) | >0.999 |
| Clarithromycin | 1 (3.2%) | 17 (13.7%) | 0.126 |
| Chloroquine | 0 (0.0%) | 1 (0.8%) | >0.999 |
| Hydroxychloroquine | 1 (3.2%) | 5 (4.0%) | >0.999 |
| Remdesivir | 0 (0.0%) | 2 (1.6%) | >0.999 |
| Anticoagulation |  |  |  |
| Profilatic |  |  |  |
| Low-molecular-weight | 16 (51.6%) | 65 (52.4%) | >0.999 |
| Non-fractionated | 11 (35.5%) | 58 (46.8%) | 0.353 |
| Fondaparinux | 0 (0.0%) | 1 (0.8%) | >0.999 |
| Therapeutic | 0 (0.0%) | 1 (0.8%) | >0.999 |
| Low-molecular-weight | 5 (16.1%) | 8 (6.5%) | 0.138 |
| Non-fractionated | 1 (3.2%) | 5 (4.0%) | >0.999 |

During hospitalization, 72 (46.5%) of patients required admission to the intensive care unit, and among them 55 (35.4%) needed mechanical ventilation and 26 (16.8%) substitutive renal therapy. Overall, there were no differences in in terms of clinical evolution and outcomes (Table 5).

**Table 5.** Clinical outcomes.

|  | **CD patients (n=31)** |  | **Control patients (n=124)** |  | p-value |
| --- | --- | --- | --- | --- | --- |
|  | Frequency (%)<br>or median<br>(IQR) | Valid cases | Frequency (%)<br>or median<br>(IQR) | Valid cases |  |
| Length of stay | 8.0 (4.5, 13.5) | 31 | 10.0 (7.0, 17.0) | 124 | 0.220 |
| Admission to ICU | 16 (51.6%) | 31 | 56 (45.2%) | 124 | 0.658 |
| Time from admission<br>to ICU (days) | 1.0 (0.0, 2.0) | 16 | 0.5 (0.0, 2.0) | 56 | 0.891 |
| Days in ICU | 6.0 (2.0, 11.2) | 16 | 7.5 (4.0, 14.0) | 56 | 0.352 |
| Thromboembolic events | 0 (0.0%) | 31 | 7 (5.6%) | 124 | 0.346 |
| Mechanical ventilation | 10 (32.3%) | 31 | 45 (36.3%) | 124 | 0.834 |
| Acute kidney injury | 9 (37.5%) | 24 | 45 (41.7%) | 108 | 0.884 |
| RRT | 5 (16.1%) | 31 | 21 (16.9%) | 124 | >0.999 |
| Sepsis | 6 (19.4%) | 31 | 24 (19.4%) | 124 | >0.999 |
| Nosocomial infection | 3 (9.7%) | 31 | 24 (19.4%) | 124 | 0.314 |
| Acute heart failure | 2 (6.5%) | 31 | 5 (4.0%) | 124 | 0.628 |
| Acute respiratory distress | 9 (29.0%) | 31 | 44 (35.5%) | 124 | 0.641 |
| Death | 10 (32.3%) | 31 | 38 (30.6%) | 124 | >0.999 |

ICU: intensive care unit; IQR: interquartile range; RRT: renal replacement therapy

## Discussion

We described a cohort of CD patients infected with SARS-COV-2 and admitted in hospitals belonging to a large Brazilian COVID-19 Registry project. Overall, CD patients had similar clinical characteristics and outcomes to non-CD controls, matched by age, sex, hypertension, DM and hospital, except from a higher prevalence of atrial fibrillation and chronic heart failure, and lower C-reactive protein levels.

Due to the potential cardiac involvement, and the higher procoagulant state, *T*.*cruzi* and SARS-COV-2 coinfection has been postulated as condition for myocardial damage, depression of ventricular function, increased arrhythmogenic state, thromboembolism risk, and ultimately a worst prognosis.[17–19] However, it was only a hypothesis and no previous study has tested it using patient data. Despite the limited number of patients with CD (31) our study refuted did not confirm the hypothesis. We did not find any significant difference or even a trend of worse clinical outcomes in CD patients, even with a higher frequency of atrial fibrillation and heart failure in the CD group.

Current data demonstrates that SARS-CoV-2 infection induces immune dysfunction, widespread endothelial injury, complement-associated coagulopathy and systemic microangiopathy. [20] By the other hand, *T. cruz*i infection is associated with an upregulated procoagulant activity in plasma. Therefore, it could be expected a greater risk of thromboembolic manifestations. In our cohort the overall thrombosis event was 4.5% (7 out of 155), all of them were in the control group. Noteworthy that, the great majority of patients (91%) were treated with oral anticoagulants because its underlying disease or received any kind of prophylactic heparin when admitted to the hospital, as recommended by national and international guidelines for the management of in-hospital COVID-19 patients. [21,22]

The lower median C-reactive level in CD patients was an unexpected finding. We hypothesize that CD patients, as they already have an active chronic inflammatory and immune response response triggered by *T*.*cruzi* infection, might have a lower risk of unregulated inflammatory response to COVID-19. [23]Therefore, what could have been a factor for worse prognosis, due to a higher frequency of associated heart failure and atrial fibrillation and the CD itself, could be equilibrated by a controlled inflammatory response. This is only a hypothesis, that merits consideration for future studies. If proved correct, it may add to the knowledge of understating how to prevent the unregulated inflammatory response in COVID-19.

It is also interesting to discuss the influence that the use of anticoagulants in full doses may have had on the outcomes of patients with CD and COVID-19. The higher prevalence of atrial fibrillation in those patients may had led to a higher frequency of use of therapeutic dosage anticoagulants (19.3% vs. 10.5%), which did not reach statistical significance due to the sample size. The best strategy to be used - prophylactic or therapeutic heparin doses - in patients with moderate to severe COVID-19 is not yet defined, and it has been hypothesized that therapeutic anticoagulation (full dose heparin) is associated with decreased in-hospital mortality in patients with moderate COVID-19, but not in patients with severe COVID-19.

It is known the effect of immunosuppressant drugs and the risk of reactivation of CD. In the case of corticosteroids, immunosuppressive doses have not been associated with higher rates of reactivation of CD, although is controversial due to the lack of supporting evidence. [24,25] Tocilizumab, a cytokine inhibitor (recombinant humanized monoclonal antibody with an antagonist effect on the IL-6 receptor), combined with another immunosuppressant agents have been suggested to be associated with the reactivation of latent infections, including parasites.

Two published case reports of Strongyloides Hyperinfection Syndrome in COVID-19 patients immunosuppressed with dexamethasone and tocilizumab, have been recently published. [26,27] To date, no cases of CD reactivation have been published, but at least, there is a concern that COVID-19 disease therapeutics could potentially trigger reactivation of CD. This merits further investigation and until definitive evidence is published, it should be a cause of concern in decision making, when prescribing immunosuppressors in these patients.

The fact that the majority of CD patients were admitted to public hospitals (81.8%) is an indicator that CD disproportionally affects people from lower income background. In a previous multivariate analysis, we demonstrated that despite being admitted to public hospitals patients do not have worse prognosis than patients admitted to private ones.[5]

This study has limitations. In addition to the retrospective design, subject to the drawbacks of a patient records review, the number of CD was low. However, it is the largest series published to date. Due to the pragmatic study design, laboratory and imaging tests were performed at the discretion of the treating physician. Despite the limited representativity of radiologic, tomographic and electrocardiographic analysis, no patient performed echocardiogram during hospital admission.

## Conclusions

Although coinfection by *Trypanosoma cruzi* and SARS-COV-2 may pose a risk of complications and therefore a worse prognosis, in our series we did not find significant differences in terms of clinical presentation and outcomes of patients with CD compared to controls, despite a higher frequency of chronic heart failure and atrial fibrillation at baseline. We observed lower C-reactive protein levels in CD when compared to controls, and this merits further investigation.

## Data Availability

Data are available upon reasonable request.

## Acknowlegments

We would like to thank the hospitals which are part of this collaboration, for supporting this Project. Especifically for this analysis, we thank the hospitals: Hospital Bruno Born; Hospital das Clínicas da UFMG; Hospital de Clínicas de Porto Alegre; Hospital Eduardo de Menezes; Hospital João XXIII; Hospital Julia Kubitschek; Hospital Metropolitano Dr. Célio de Castro; Hospital Nossa Senhora da Conceição; Hospital Regional Antônio Dias; Hospital Risoleta Tolentino Neves; Hospital Unimed-BH.

We also thank all the clinical staff at those hospitals, who cared for the patients, and all undergraduate students who helped with data collection.

## Data availability statement

Data are available upon reasonable request.

## Transparency declaration

The lead authors (MSM, IM and MCP) affirm that the manuscript is an honest, accurate, and transparent account of the study being reported; that no important aspects of the study have been omitted; and that any discrepancies from the study as originally planned (and, if relevant, registered) have been explained.

## Author contribution

Substantial contributions to the conception or design of the work: MSM, IMR, IJRO,MCP, MCPN.

Substantial contributions to the acquisition, analysis, or interpretation of data for the work: IMR, MSM, LMO, MCP, RTS, MHCG, IJRO, LSM, RLRC, AGSN, ANRMB, ANBAS, AACM, BLF, CAC, FDAR, FA, FAB, GFN, HD, HCG, JCA, LBM, LBZ, LFA, LK, LCC, LEAS, MASC, MAPF, NCSS, NRO, SCF, SJTSL, VBS, VRBC, YCRFABM, RM, MCPN

Drafted the work: IMR, MSM, MCP, MCPN.

Revised the manuscript critically for important intellectual content: all authors. Final approval of the version to be published: all authors.

Drafted the work: IM, MSM, MCP, MCPN.

Revised the manuscript critically for important intellectual content: all authors.

Final approval of the version to be published: all authors.

Agreement to be accountable for all aspects of the work in ensuring that questions related to the accuracy or integrity of any part of the work are appropriately investigated and resolved: MSM and MCP.

## Figure

**Table S1.** Laboratory parameters of the study population at baseline.

|  | CD patients (n=31) |  | Controls (n=124) |  | p-value |
| --- | --- | --- | --- | --- | --- |
|  | Frequency (%)<br>or median (IQR) | Valid cases | Frequency (%)<br>or median (IQR) | Valid cases |  |
| Hemoglobin (g/dL) | 12.8 (12.1, 14.1) | 28 | 12.9 (11.8, 14.0) | 120 | 0.849 |
| White blood cell count (x10<sup>9</sup>/L) | 6.3 (4.7, 8.0) | 28 | 7.0 (5.1, 10.0) | 120 | 0.205 |
| Neutrophils (x10<sup>9</sup>/L) | 4,142.5<br>(3,287.5, 6,102.2) | 28 | 5,548.5<br>(3,528.8, 7,597.5) | 118 | 0.154 |
| Lymphocytes (x10<sup>9</sup>/L) | 960.0<br>(609.8, 1,443.8) | 28 | 1,001.0<br>(688.2, 1,291.5) | 116 | 0.824 |
| Platelets (x10<sup>9</sup>/L) | 221.5<br>(147.3, 319.8) | 28 | 194.2<br>(138.3, 272.5) | 119 | 0.186 |
| Albumin (g/dL) | 3.3 (2.9, 3.7) | 13 | 3.2 (2.9, 3.6) | 50 | 0.792 |
| AST (U/L) | 45.0 (28.8, 50.5) | 20 | 46.0 (33.0, 69.0) | 79 | 0.351 |
| ALT (U/L) | 26.5 (14.0, 38.2) | 20 | 30.0 (21.8, 47.0) | 80 | 0.154 |
| Calcium (mmol/L) | 1.1 (1.0, 1.2) | 10 | 1.1 (1.1, 1.1) | 39 | 0.302 |
| Creatinine (mg/dL) | 1.0 (0.9, 1.2) | 27 | 0.9 (0.7, 1.3) | 117 | 0.546 |
| CPK (U/L) | 87.5 (64.0, 183.0) | 10 | 107.0 (47.0, 306.0) | 41 | 0.553 |
| C-reactive protein (mg/dL) | 55.5 (35.7, 85.0) | 28 | 94.3 (50.7, 167.5) | 115 | **0.013** |
| D-dimer (ng/mL) | 1.9 (1.1, 600.0) | 17 | 4.0 (1.9, 1,765.0) | 68 | 0.106 |
| LDH (U/L) | 435.0<br>(311.0, 471.5) | 19 | 381.5<br>(285.0, 515.2) | 76 | 0.930 |
| Lactate (mmol/L) |  | 31 |  | 123 | 0.863 |
| Potassium (mmol/L) | 4.3 (3.8, 4.7) | 27 | 4.3 (3.9, 4.7) | 116 | 0.782 |
| aPTT (seconds) | 1.1 (1.0, 1.4) | 22 | 1.0 (0.9, 1.1) | 87 | **0.004** |
| INR | 1.2 (1.1, 1.6) | 23 | 1.1 (1.0, 1.2) | 89 | **0.018** |
| Sodium (mmol/L) | 137.0<br>(133.9, 139.8) | 26 | 137.0<br>(133.6, 141.0) | 115 | 0.856 |
| Total bilirubin (mg/dL) | 0.4 (0.3, 0.6) | 18 | 0.5 (0.3, 0.7) | 83 | 0.866 |
| Troponin (ng/mL) | 0.6 (0.5, 3.5) | 9 | 0.6 (0.1, 1.0) | 40 | 0.366 |
| Urea (mg/dL) | 40.5 (32.8, 59.8) | 28 | 42.0 (31.0, 67.2) | 118 | 0.673 |

**Table S2.** Radiological characteristics of the study population at baseline.

|  | **CD patients**<br>**(n=31)** | **Controls**<br>**(n=124)** | **p-value** |
| --- | --- | --- | --- |
| **Chest X-ray at admission** | **19 (61.3%)** | **91 (73.4%)** | 0.269 |
| Normal | 3 (15.8%) | 19 (20.9%) | 0.760 |
| Atelectasis | 0 (0.0%) | 2 (2.2%) | >0.999 |
| Consolidation | 6 (31.6%) | 15 (16.5%) | 0.195 |
| Diffuse interstitial infiltrate | 9 (47.4%) | 52 (57.1%) | 0.599 |
| Focal interstitial infiltrate | 1 (5.3%) | 6 (6.6%) | >0.999 |
| Bilateral ground glass opacity | 6 (31.6%) | 17 (18.7%) | 0.223 |
| **Chest X-ray at follow up** | **11 (35.5%)** | **58 (46.8%)** | 0.353 |
| Normal | 3 (27.3%) | 9 (15.5%) | 0.390 |
| Atelectasis | 0 (0.0%) | 2 (3.4%) | >0.999 |
| Consolidation | 2 (18.2%) | 17 (29.3%) | 0.715 |
| Diffuse interstitial infiltrate | 5 (45.5%) | 33 (56.9%) | 0.525 |
| Focal interstitial infiltrate | 0 (0.0%) | 4 (6.9%) | >0.999 |
| Bilateral ground glass opacity | 6 (54.5%) | 14 (24.1%) | 0.067 |
| Radiological progression | 3 (27.3%) | 11 (19.0%) | 0.683 |
| **Chest CT at admission** | **8 (25.8%)** | **48 (38.7%)** | 0.259 |
| Normal | 1 (12.5%) | 3 (6.2%) | 0.470 |
| Atelectasis | 2 (25.0%) | 3 (6.2%) | 0.144 |
| Consolidation | 2 (25.0%) | 9 (18.8%) | 0.649 |
| Pleural effusion | 0 (0.0%) | 2 (4.2%) | >0.999 |
| Bilateral ground glass opacity | 6 (75.0%) | 40 (83.3%) | 0.623 |
| Peripheral ground glass opacity | 5 (62.5%) | 25 (52.1%) | 0.712 |
| Crazy-paving pattern | 0 (0.0%) | 6 (12.5%) | 0.578 |
| **Chest CT at follow up** | **5 (16.1%)** | **31 (25.0%)** | 0.419 |
| Normal | 0 (0.0%) | 5 (16.1%) | >0.999 |
| Atelectasis | 1 (20.0%) | 2 (6.5%) | 0.370 |
| Consolidation | 0 (0.0%) | 7 (22.6%) | 0.559 |
| Pleural effusion | 3 (60.0%) | 2 (6.5%) | **0.013** |
| Unilateral ground glass opacity | 0 (0.0%) | 2 (6.5%) | >0.999 |
| Bilateral ground glass opacity | 3 (60.0%) | 19 (61.3%) | >0.999 |
| Peripheral ground glass opacity | 3 (60.0%) | 19 (61.3%) | >0.999 |
| Crazy-paving pattern | 2 (40.0%) | 6 (19.4%) | 0.305 |
| Progression<sup>*</sup> | 5 (100.0%) | 30 (96.8%) | >0.999 |

\* Normal at admission and abnormal at follow-up
CT: computer tomography.

